# Shared genetic etiology between idiopathic pulmonary fibrosis and COVID-19 severity

**DOI:** 10.1101/2020.12.15.20248279

**Authors:** João Fadista, Luke M. Kraven, Juha Karjalainen, Shea J. Andrews, Frank Geller, The COVID-19 Host Genetics Initiative, J Kenneth Baillie, Louise V. Wain, R. Gisli Jenkins, Bjarke Feenstra

## Abstract

**Background:** Idiopathic pulmonary fibrosis (IPF) is a complex lung disease, characterized by progressive lung scarring. Severe COVID-19 is associated with substantial pneumonitis and has a number of shared major risk factors with IPF. This study aimed to determine the genetic correlation between IPF and severe COVID-19 and assess a potential causal role of genetically increased risk of IPF on COVID-19 severity.

**Methods:** We performed a Mendelian randomisation (MR) study for IPF causality in COVID-19. Genetic variants associated with IPF susceptibility (P<5×10^−8^) in previous genome-wide association studies (GWAS) were used as instrumental variables (IVs). Effect estimates of those IVs on COVID-19 severity were gathered from the GWAS meta-analysis by the COVID-19 Host Genetics Initiative. The genetic correlation between IPF and COVID-19 severity was estimated with linkage disequilibrium (LD) score regression.

**Findings:** We detected a positive genetic correlation of IPF with COVID-19 severity (rg=0.31 [95% CI 0.04-0.57], P = 0.023). The MR estimates for severe COVID-19 did not reveal any genetic association (OR 1.05, [95% CI 0.92-1.20], P = 0.43). However, outlier analysis revealed that the IPF risk allele rs35705950 at *MUC5B* had a different effect compared with the other variants. When rs35705950 was excluded, MR results provided evidence that genetically increased risk of IPF has a causal effect on COVID-19 severity (OR 1.21, [95% CI 1.06-1.38], P = 4.24×10^−3^). Furthermore, the IPF risk-allele at *MUC5B* showed an apparent protective effect against COVID-19 hospitalization only in older adults (OR 0.86, [95% CI 0.73-1.00], P = 2.99×10^−2^).

**Interpretation:** The strongest genetic determinant of IPF, rs35705950 at *MUC5B*, seems to confer protection against COVID-19, whereas the combined effect of all other IPF risk loci seem to confer risk of COVID-19 severity. The observed effect of rs35705950 could either be due to protective effects of mucin over-production on the airways or a consequence of selection bias due to a patient group that is heavily enriched for the rs35705950 T undertaking strict self-isolation. Due to the diverse impact of IPF causal variants on SARS-CoV-2 infection, further investigation is needed to address this apparent paradox between variance at *MUC5B* and other IPF genetic risk factors.

**Funding:** Novo Nordisk Foundation and Oak Foundation.

## INTRODUCTION

Since the emergence of a novel severe acute respiratory syndrome (SARS) coronavirus 2(SARS-CoV-2) in Wuhan, China in December 2019, there have been more than 70 million confirmed cases and over 1.6 million deaths worldwide^1^. SARS-CoV-2 infection, which causes coronavirus disease 2019 (COVID-19), ranges from asymptomatic to severe disease needing ICU admission and mechanical ventilation^2^. Infection estimates may vary considerably within populations due to the frequency of asymptomatic disease and inherent risk factors for symptomatic disease as well as public health protection policies^3^. It is estimated that about 45% of those infected are asymptomatic, while up to 10% require hospitalization^4,5^. Severe disease, which occurs in up to 20% of hospitalized patients, is associated with a high mortality rate^6^.

Idiopathic pulmonary fibrosis (IPF) is a complex lung disease characterized by progressive lung scarring caused by damage to the alveolar epithelium followed by an abnormal wound-healing response causing deposition of dense fibrotic tissue, which ultimately leads to loss of lung function and death through respiratory failure^7^. Moreover, despite the drugs pirfenidone and nintedanib being approved for IPF treatment, there is still no cure as these drugs only slow disease progression, and half of IPF patients die within 3 to 5 years after diagnosis^7,8^. IPF is influenced by both genetic and environmental factors. Previous genome-wide association studies (GWAS) have revealed common genetic variants associated with IPF^9,10^, with the largest GWAS of IPF to date detecting 14 loci^9^. The most strongly associated IPF variant^9,11^, rs35705950, has its risk allele (T) associated with a five-fold increase in disease risk^9^ and over expression of mucin 5B in small-airway epithelial cells^12,13^.

COVID-19 and IPF both begin with lung injury, and their most severe consequences are seen in elderly males, with male IPF patients showing a high risk of COVID-19 mortality^14,15^. Thus, it is plausible that there are shared pathogenic mechanisms between severe COVID-19 and IPF, which may relate to an underlying shared genetic etiology. Should there be shared genetic and pathological mechanisms, this would provide some rationale for investigating whether repurposing of anti-fibrotic therapy could be a treatment strategy for patients with COVID-19.

This study uses the genetic determinants of IPF, estimated from IPF GWAS summary statistics^9,10^ to perform two-sample Mendelian randomization analysis^16^ to assess whether a causal relationship between genetically mediated IPF risk and COVID-19 severity is plausible^17^. In addition, we analyze common genetic variation across the entire genome to estimate the genetic correlation between IPF^9^ and COVID-19 severity^17^.

## METHODS

### Study population

We extracted association summary statistics from the largest meta-analysis GWAS of IPF to date (4,124 cases and 20,465 controls)^9^ as well as the only exome-wide association study (ExWAS) of IPF using whole genome and whole exome sequenced samples (752 cases and 119,055 controls)^10^. All IPF cases and controls were of genetically determined European ancestries. The summary statistics for the outcome of COVID-19 severity was extracted from the COVID-19 Host Genetics Initiative (HGI) GWAS meta-analysis^17^, available at https://www.covid19hg.org/results/. These COVID-19 HGI summary statistics are from the fourth round of GWAS meta-analysis, publicly available since October 20th, 2020, but without the 23andMe cohort. The COVID-19 HGI is an international collaborative effort that aims to study the genetic determinants of COVID-19 susceptibility, hospitalization and severity. The COVID-19 HGI has gathered clinical and genetic data and performed GWAS meta-analysis of multiple cohorts with a fixed effects inverse variance weighting. The analysis was adjusted for age, age^2^, sex, age*sex, genetic ancestry principal components and other study-specific covariates. An allele frequency of 0.001 and an imputation info score of 0.6 was applied to each study before meta-analysis. Severe cases were defined as hospitalized patients with confirmed COVID-19 by RNA PCR, serologic testing, or physician diagnosis that had very severe respiratory complications (N=4,336). Population controls were defined as individuals who tested negative for COVID-19, were never tested, or had an unknown testing status (N=623,902). About 89.6% of COVID-19 severity cases and 99.9% of controls were of European ancestries. Data was also extracted from GWAS of COVID-19 hospitalization (6,406 cases and 902,088 controls) and COVID-19 susceptibility (14,134 cases and 1,284,876 controls), restricted to European ancestry individuals. GWAS using non-hospitalized COVID-19 cases as controls for hospitalized COVID-19 cases was also analyzed (2,430 cases and 8,478 controls). Similarly, GWAS using lab and/or self-reported negative COVID-19 individuals as controls for COVID-19 positive cases was also used (24,057 cases and 218,062 controls).

### Genetic instrument variants

We used the 15 independent genetic variants associated with IPF at genome wide significance (P<5×10^−8^) as instrumental variables (Supplementary Table 1). For the only genetic variant out of the 15 that was not represented in the COVID-19 severity outcome GWAS^17^, we selected the next best available genetic variant based on posterior probability from IPF credible sets^9^, while having an LD r^2^>0.9 with the index variant using the CEU European sub-cohort of the 1000 Genomes Phase 3 dataset (original SNP is rs2077551; proxy SNP is rs17652520) (Supplementary Table 1). Of the 15 genetic instruments, rs35705950 at the *MUC5B* gene locus explains 5.9-9.4% of IPF liability in the general population, while the remaining 14 loci collectively explain up to 3% of IPF liability in the general population^11^.

**Table 1.** MR effect estimates of IVW, Weighted median and MR-Egger methods on the causal association of IPF with COVID-19 severity. Effect estimates are for all 15 IVs, 14 IVs without the rs35705950 at the *MUC5B* locus, and the 5 non-MUC5B IVs that do not associate with any other phenotype (Methods). IVs, genetic instrumental variables. OR, odds-ratio. CI Lower, lower confidence interval estimate. CI Upper, upper confidence interval estimate. P, p-value.

| Analysis | OR | 95% CI Lower | 95% CI Upper | P | Method |
| --- | --- | --- | --- | --- | --- |
| All 15 IVs | 1.054 | 0.924 | 1.203 | 4.31E-01 | IVW |
|  | 1.042 | 0.937 | 1.158 | 4.49E-01 | Weighted median |
|  | 0.831 | 0.680 | 1.017 | 7.25E-02 | MR-Egger |
| 14 IVs<br>(no *MUC5B*) | 1.210 | 1.062 | 1.379 | 4.24E-03 | IVW |
|  | 1.188 | 1.074 | 1.315 | 8.62E-04 | Weighted median |
|  | 1.296 | 0.669 | 2.510 | 4.43E-01 | MR-Egger |
| 5 IVs<br>(no other pheno) | 1.402 | 1.158 | 1.697 | 5.34E-04 | IVW |
|  | 1.349 | 1.164 | 1.565 | 7.30E-05 | Weighted median |
|  | 1.233 | 0.442 | 3.439 | 6.89E-01 | MR-Egger |
| 11 IVs<br>(no MR-PRESSO outliers) | 1.194 | 1.106 | 1.290 | 5.48E-06 | IVW |
|  | 1.107 | 0.998 | 1.227 | 5.54E-02 | Weighted median |
|  | 1.290 | 0.907 | 1.834 | 1.56E-01 | MR-Egger |

### Mendelian Randomization analysis

To investigate causality between IPF and COVID-19 severity, two-sample Mendelian randomization (MR) analysis was performed using the random-effects inverse-variance weighted method (IVW)^18^, implemented in the R (version 3.6.1)^19^ package *MendelianRandomization* (version 0.5.0). MR relies on three main assumptions: (1) the instrumental variables must be strongly associated with the exposure, (2) must not be associated with factors that confound the relationship between exposure and outcome, and (3) can only be associated with the outcome through the exposure. Sensitivity analysis was performed with the weighted median^20^ and MR-Egger methods^21^, which are less powerful than the IVW method if all MR assumptions hold, but are more robust to invalid instruments and horizontal pleiotropy^22^. The random-effects IVW method^18^, which assumes that all genetic variants are valid instruments, regresses the effect sizes of variant COVID-19 severity associations against effect sizes of the variant IPF associations, assuming that the strength of the association of the genetic instruments with IPF is not correlated with the magnitude of the pleiotropic effects or that the pleiotropic effects have an average value of zero. On the contrary, since the weighted median method uses the median instrumental variable from all variants, it is robust to pleiotropy when <50% of the genetic variants are invalid^20^. By using a weighted regression with an unconstrained intercept, MR-Egger^21^ does also not assume that all variants are valid instruments. If the MR-Egger intercept term differs significantly from zero, then the genetic variants are not all valid. An IVW leave-one-out analysis, implemented in the *MendelianRandomization* R package was also performed to determine whether any outliers could bias the overall causal estimate.

Pleiotropy against other phenotypes was also determined through PheWAS lookups of each genetic instrumental variable against Phenoscanner^23^ and GeneATLAS^24^. Association results were deemed significant at P<1×10^−5^, and restricted to non-cancer diseases, non-lung and non-blood trait phenotypes. MR-PRESSO outlier test^25^, implemented in the *MRPRESSO* (version 1.0) R package^19^ was also performed to detect horizontal pleiotropic IVs.

### Age-stratified COVID-19 severity GWAS

To detect possible age-related effects of the IPF genetic instruments on COVID-19 severity, two GWAS of COVID-19 severity were performed for individuals greater and lower than 60 years of age, respectively.

### Genetic correlation

Analysis of genetic correlation between IPF and COVID-19 severity was performed using the LD score regression method^26,27^ applied to GWAS summary statistics of both diseases^9,17^ using only variants with MAF>1% in both GWAS that were present in the HapMap3 recommended SNP list^26^.

## RESULTS

### Genetic correlation of IPF with COVID-19 severity

To quantify the shared genetic etiology of IPF with COVID-19 severity, we used the LD score regression method^26,27^ and detected a positive genetic correlation of IPF susceptibility with COVID-19 severity (rg=0.31 [95% CI 0.04-0.57], P = 0.023), which suggested a shared genetic etiology. A positive genetic correlation with COVID-19 hospitalization (rg=0.31 [95% CI 0.02-0.63], P = 0.035) was also detected, but at low statistical significance and consequent wider confidence intervals. The genetic correlation between IPF risk and COVID-19 susceptibility was positive, though not statistically significant (rg=0.25 [95% CI -0.13-0.62], P = 0.193).

### Mendelian Randomization analysis of COVID-19 severity

To determine whether the genetic correlation results have a known causal component, a two-sample MR analysis was performed to test the causal effect of IPF risk genes on COVID-19 severity. Genetically increased IPF risk was not associated with a higher risk of COVID-19 severity when compared to population controls using random-effects inverse variance weighted method (IVW)^18^ on all 15 genetic instruments (OR 1.05, [95% CI 0.92-1.20], P = 0.43) (Table 1). However, a highly significant heterogeneity of effects (P=3.30×10^−16^) prompted us to do IVW leave-one-out analysis to detect possible outlying genetic variants. As seen in Figure 1, rs35705950 at the *MUC5B* gene locus appeared to be an outlier. Re-running the IVW MR analysis on the 14 non-*MUC5B* genetic instruments, we detected that a genetically increased IPF risk was associated with a higher risk of COVID-19 severity (OR 1.21, [95% CI 1.06-1.38], P = 4.24×10^−3^) (Table 1) (Figure 2). Sensitivity analysis with the weighted median method^20^ detected similar significant effect estimates (OR 1.19, [95% CI 1.07-1.32], P = 8.62×10^−4^). Despite the reduced power of MR-Egger^21^ compared to the IVW method^18^, we detected consistent effect estimates for increased IPF risk and COVID-19 severity, albeit with broad confidence intervals (OR 1.30, [95% CI 0.67-2.51], P = 0.44) (Table 1). Moreover, the MR-Egger intercept test indicated the absence of directional pleiotropy (P = 0.84).

**Figure 1.**
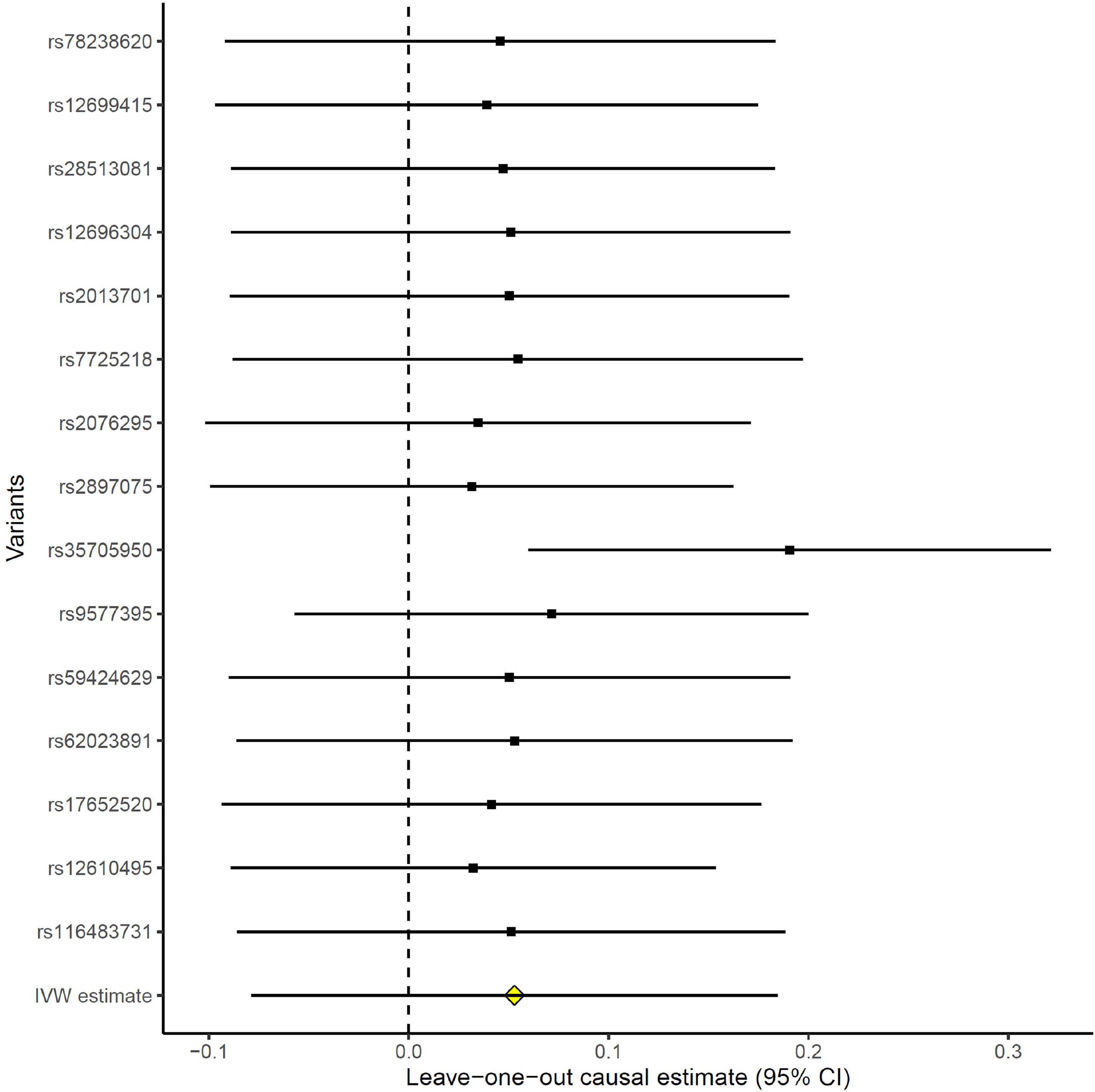
Forest plot of IVW causal estimates, omitting each variant in turn. The estimate with the first labelled SNP includes all variants except the labelled variant, and so on. The IVW estimate including all variants (“IVW estimate”) is also provided for reference. Estimates are in ln(OR).

**Figure 2.**
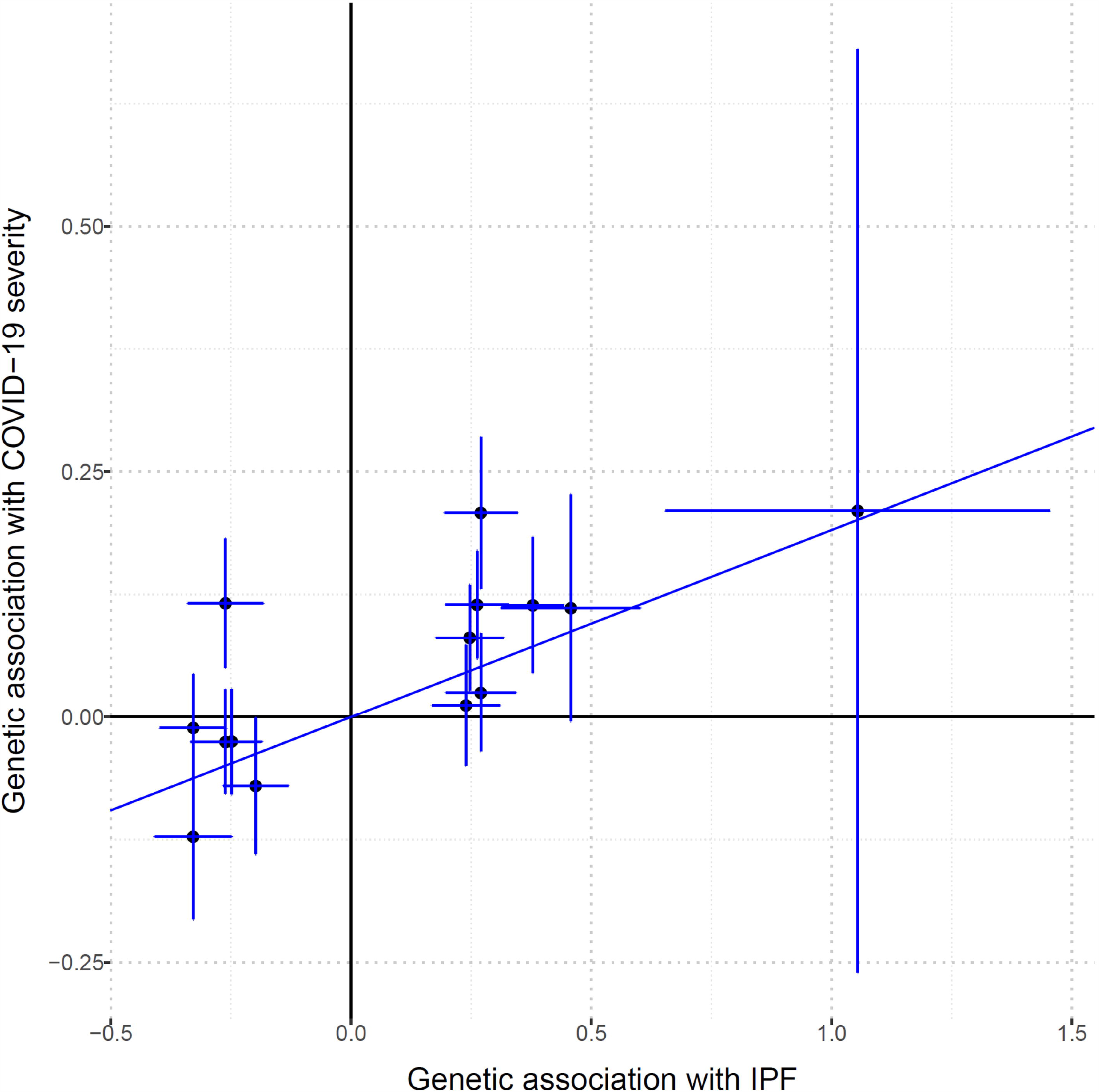
Genetic association estimates of the 14 non-*MUC5B* instrumental variables not detected to be outliers. Horizontal error bars regards standard errors of IPF estimates, while vertical error bars regards standard errors of COVID-19 severity estimates. The line represents the IVW causal estimate of IPF on COVID-19 severity. IPF, idiopathic pulmonary fibrosis. Estimates are in ln(OR).

We also used Phenoscanner^23^ and GeneATLAS^24^ to detect if any of the genetic instrumental variables used in this MR study were associated with any other phenotypes. Eight out of the 14 non-MUC5B IVs were associated with at least one other non-cancer, non-lung and non-blood disease/trait. Those are rs12699415, rs28513081, rs12696304, rs7725218, rs9577395, rs59424629, rs62023891 and rs17652520 (Supplementary Table 1). Removing these 8 SNPs left us with 5 non-*MUC5B* SNPs (Supplementary Table 1). While the confidence intervals widened, the MR effect estimates restricting the analysis to these 5 SNPs remained consistent for IVW (OR 1.40, [95% CI 1.21-1.59], P = 5.34×10^−4^), weighted median (OR 1.35, [95% CI 1.20-1.50], P = 7.30×10^−5^) and MR-Egger (OR 1.23, [95% CI 0.21-2.26], P = 0.69) (Table 1). We also used MR-PRESSO^25^ to further detect horizontal pleiotropic outlier IVs. Four out of the 14 IVs were detected as outliers (rs35705950 at *MUC5B*, rs2897075, rs9577395 and rs12610495) (Supplementary Table 1). Removing these 4 SNPs left us with 11 SNPs. MR effect estimates using these 11 IVs for IVW (OR 1.19, [95% CI 1.11-1.29], P = 5.48×10^−6^), weighted median (OR 1.11, [95% CI 1.00-1.23], P = 5.54×10^−2^) and MR-Egger (OR 1.29, [95% CI 0.91-1.83], P = 0.16) also remained consistent, i.e. increased IPF genetic risk associated with increased COVID-19 severity (Table 1).

### *MUC5B* vs. COVID-19 with different control groups and in an age-stratified analysis

Since rs35705950 at the MUC5B gene locus was an outlier in the MR analysis, we assessed its association with COVID-19 susceptibility, hospitalization and severity. While the rs35705950 IPF risk allele T seems to be protective for COVID-19, its effect estimates and significance decrease as the case inclusion criteria expand to include less severe cases (Table 2). Using non-hospitalized COVID-19 cases as controls, rather than population controls, compared with hospitalized COVID-19 cases further decreased the protective effect size of the rs35705950 T allele (Figure 3). Similarly, using lab/self-reported negative individuals as controls (instead of population controls) against COVID-19 positive cases also decreased the protective effect size of the rs35705950 T allele (Figure 3). Furthermore, age stratified analysis for COVID-19 hospitalization and susceptibility showed that while the protective effect for hospitalisation was similar for both age groups, albeit non-significant in under 60s (over 60: hospitalization OR 0.86, [95% CI 0.73-1.00], P = 2.99×10^−2^; under 60: hospitalization OR 0.85, [95% CI 0.64-1.06], P = 0.14), the effect on susceptibility was reduced to the null in the under 60s (over 60: susceptibility OR 0.87, [95% CI 0.77-0.98], P = 9.48×10^−3^; under 60: susceptibility OR 0.99, [95% CI 0.90-1.09], P = 0.91) (Table 2).

**Table 2.** Effect estimates of rs35705950 at the *MUC5B* locus on COVID-19 severity (4,336 cases & 623,902 controls), hospitalization (6,406 cases & 902,088 controls) and susceptibility (14,134 cases & 1,284,876 controls). All, all the cohort. >60, sub-cohort of individuals older than 60 years. <60, sub-cohort of individuals younger than 60 years. OR, odds-ratio of the IPF T risk allele. CI Lower, lower confidence interval estimate. CI Upper, upper confidence interval estimate. P, p-value. AF, allele frequency of rs35705950 T allele. N, sample size.

| Age | COVID-19 | OR | 95% CI Lower | 95% CI Upper | P | AF | N |
| --- | --- | --- | --- | --- | --- | --- | --- |
| All | Severity | 0.78 | 0.66 | 0.90 | 6.50E-05 | 0.108 | 628,238 |
|  | Hospitalization | 0.83 | 0.74 | 0.92 | 3.77E-05 | 0.112 | 908,494 |
|  | Susceptibility | 0.93 | 0.88 | 0.99 | 9.18E-03 | 0.116 | 1,299,010 |
| >60 | Hospitalization | 0.86 | 0.73 | 1.00 | 2.99E-02 | 0.109 | 417,774 |
|  | Susceptibility | 0.87 | 0.77 | 0.98 | 9.48E-03 | 0.110 | 470,162 |
| <60 | Hospitalization | 0.85 | 0.64 | 1.06 | 1.42E-01 | 0.112 | 83,265 |
|  | Susceptibility | 0.99 | 0.90 | 1.09 | 9.07E-01 | 0.107 | 314,152 |

**Figure 3.**
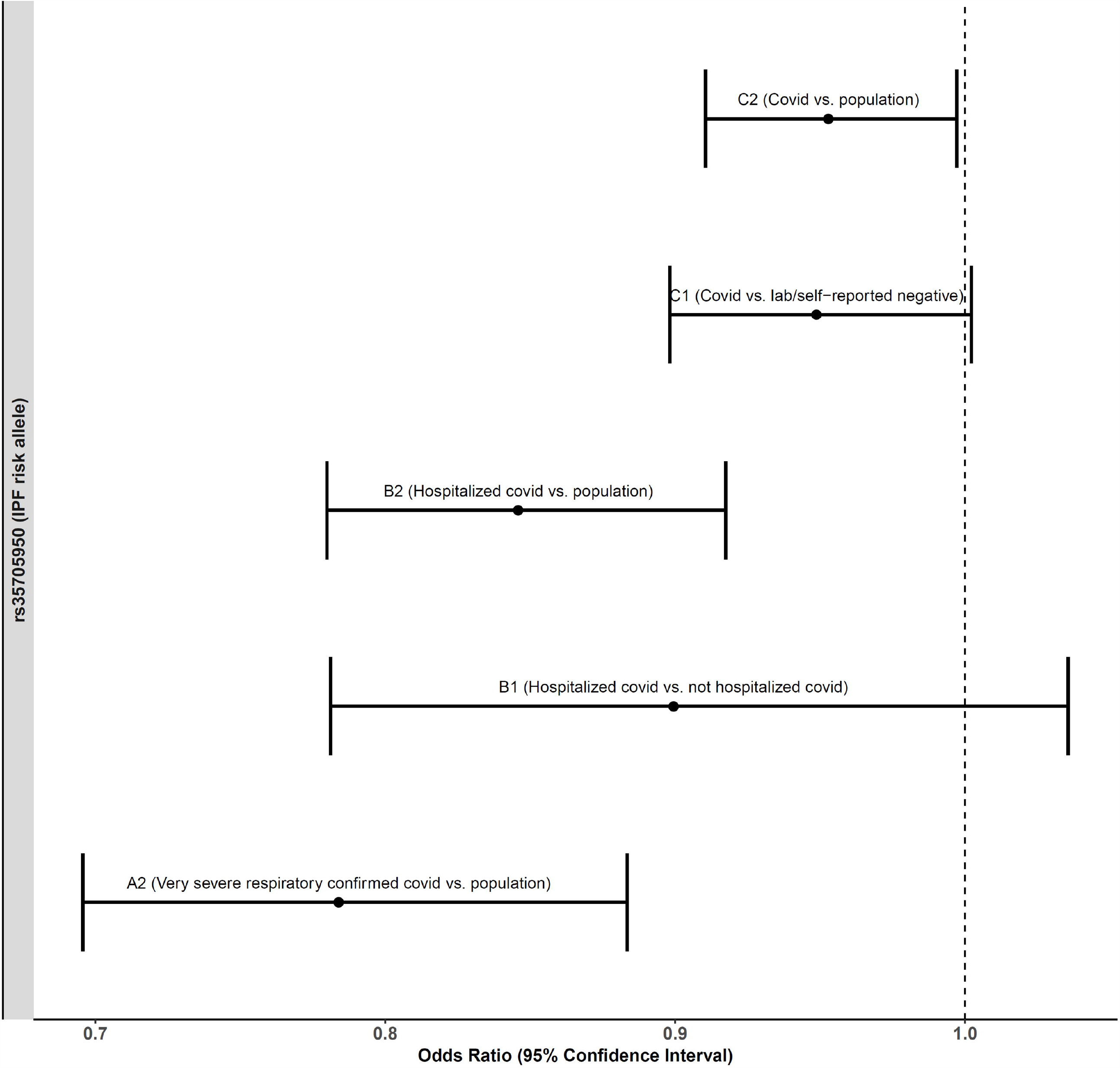
Forest plot of the effect estimates of the association of rs35705950 IPF T risk allele at the *MUC5B* locus on various COVID-19 outcomes using different control population. A2 (Very severe respiratory confirmed covid vs. population) has 4,336 cases and 623,902 controls. B1 (Hospitalized covid vs. not hospitalized covid) has 2,430 cases and 8,478 controls. B2 (Hospitalized covid vs. population) has 6,406 cases and 902,088 controls. C1 (Covid vs. lab/self-reported negative) has 24,057 cases and 218,062 controls. C2 (Covid vs. population) has 14,134 cases and 1,284,876 controls.

## DISCUSSION

Patients with Idiopathic pulmonary fibrosis (IPF) are at increased risk of COVID-19 mortality compared with the general population^14,15^. Whether the pathogenic mechanisms that lead to the development of IPF are causally related to the severity of COVID-19 is unknown and of paramount importance to inform preventive strategies and identify whether there is rationale for investigating the role of anti-fibrotic therapies in severe COVID-19^28^. Using a two-sample Mendelian randomization approach and genome-wide genetic correlation analysis, we found that overall there was a genetic correlation between IPF and COVID-19 severity, but the genetic variants associated with IPF did not confer an increased risk of severe COVID-19. However, this was driven by a single outlier variant at the *MUC5B* locus, which had an apparently protective effect on the severity of COVID-19. Removal of this outlier demonstrated that, collectively, the remaining variants associated with increased IPF risk were associated with increased risk of severe COVID-19 (Table 1). This finding supports the epidemiological studies that have reported a strong association between IPF and COVID-19 severity^14,15,29^.

It is intriguing that the most strongly associated IPF variant^9,11^, rs35705950 at the *MUC5B* gene locus, which its risk allele (T) associated with a five-fold increase in IPF risk^9^, is negatively associated with COVID-19 regardless of severity, suggesting that this IPF risk allele may protect against COVID-19. This association has been reported in a small study^30^, and a protective role for *MUC5B* in airway defense has been described^31^. The IPF risk allele (T) of the rs35705950 is associated with increased *MUC5B* expression in lung tissue^13^, which has been associated with muco-ciliary dysfunction and increased bleomycin-induced fibrosis in mice^32^.

Alternatively, this apparent protective effect of the rs35705950 T allele against COVID-19 could be the consequence of selection bias in the COVID-19 GWAS. Whilst there is expected to be minimal misclassification in the COVID-19 case definitions, the general population control groups are likely to contain individuals who have never been exposed to the virus and thus whose severity of response to the virus is, as yet, unknown. The IPF patient population will be enriched for the rs35705950 T allele but less likely to be amongst the cases due to shielding behavior. The less protective effect of the rs35705950 T allele against COVID-19 when using COVID-19 negative individuals or non-hospitalized COVID-19 individuals as controls instead of using population controls further suggests that there might be a selection bias. The strong risk effect of this variant has previously been shown to introduce a bias in survival analyses^33^. This could account for the reduced protective effect of rs35705950 T allele in the younger age group strata of the COVID-19 GWAS as IPF is predominantly a disease affecting those over 60. However, the remaining 14 IPF variants are associated with an increased risk of severe COVID-19. Both IPF and severe COVID-19 are associated with increasing age and obesity and this may suggest shared a pathogenic role for cellular senescence^34,35^ or metabolic syndrome^36^. The IPF genetic risk variant near the *DPP9* gene on chromosome 19 has recently been reported as genome-wide significantly associated with COVID-19 severity^37^. In light of our findings of shared causal genetic etiology, it suggests that some of the molecular mechanisms that lead to IPF could also be important in the response to COVID-19. If that is the case, as already hypothesized in the literature^28,38^, antifibrotic therapies used to treat IPF could have an important role in mitigating COVID-19 severity in IPF patients, and could potentially be evaluated in clinical trials to prevent the development of COVID-19 emergent pulmonary fibrosis.

Our study also has some limitations, such as the modest variance explained by the IPF genetic instruments, although within the range typical of complex traits. Nevertheless, the use of weak genetic instruments could only create biases estimates towards the null. Increased sample sizes, both from the IPF or COVID-19 GWAS could also have narrowed our confidence intervals around the true estimates, although we used the largest sample sizes for IPF and COVID-19 to date. Furthermore, MR-Egger results were not as compelling as the IVW or weighted median, suggesting that confounding factors could have biased the effect estimates. However, MR-Egger is usually considered as a sensitivity method which can also be biased in certain situations^39^. Moreover, the COVID-19 control groups, drawn from the general population, were of unknown virus exposure status, which might have further biased our causal estimates towards the null.

In summary, our study provides genetic evidence that supports shared causal genetic etiology between IPF and COVID-19 severity that could inform the design of future preventive and therapeutic strategies to treat COVID-19.

## Data Availability

Supporting data is available in Supplementary Material, on https://www.covid19hg.org/results/ and https://github.com/genomicsITER/PFgenetics.

ttps://www.covid19hg.org/results/

https://github.com/genomicsITER/PFgenetics

## DECLARATIONS

### Ethical Approval and Consent to participate

Covid-19 HGI and the IPF GWAS consortia have ethical approvals from their respective cohorts [14,15,22]. Patients or the public were not involved in the design, or conduct, or reporting, or dissemination plans of our research.

### Consent for publication

All authors consent this study for publication.

### Authors’ contributions

JF conceptualized, designed the study, coordinated data collection and carried out the initial analyses and drafted the initial manuscript. LMK, JK and SJA collected data and also carried out initial analyses. JKB, FG, and AG critically reviewed the manuscript for important intellectual content. BF, RGJ and LVW conceptualized, designed the study, coordinated and supervised data collection and analysis, and critically reviewed the manuscript for important intellectual content. All authors approved the final manuscript as submitted and agree to be accountable for all aspects of the work.

## Acknowledgements

We thank the patients, staff and investigators who contributed to the Covid-19 HGI and the IPF GWAS consortia.

